# Oxidative Stress Biomarker Profile Dynamics across Blood and Cerebrospinal Fluid

**DOI:** 10.64898/2026.06.19.26355411

**Authors:** Adrián Noriega de la Colina, Zoe Skaperda, Sokratis Charisis, Eva Ntanasi, Eirini Mamalaki, Maria Yannakoulia, Christopher Papandreou, Fotios Tekos, Dimitrios Kouretas, Nikolaos Scarmeas

## Abstract

Peripheral blood measurements dominate oxidative stress research, yet whether they reflect central nervous system (CNS) redox status remains untested in humans. We simultaneously profiled five biomarkers, total antioxidant capacity (TAC), glutathione (GSH), thiobarbituric acid-reactive substances (TBARS), ferric reducing antioxidant power (FRAP), and hydroxyl radical scavenging activity (HRSA), in paired blood and cerebrospinal fluid (CSF) from 140 adults in the ALBION cohort. Only FRAP showed a significant positive cross-compartment correlation (ρ = +0.49, FDR-p < 0.001), supporting its role as a systemic antioxidant signal. TBARS showed a significant inverse cross-compartment association (ρ = −0.20, FDR-p = 0.042), suggesting compartmental compensation in lipid peroxidation regulation rather than parallel dynamics. TAC and GSH showed no meaningful intercompartmental alignment. Individual biomarker levels were largely stable across the 40-85 year age range in both compartments, suggesting that age effects operate through coordinated latent networks rather than single-marker trajectories. Principal component extraction with varimax rotation identified four latent factors explaining 66.6% of total variance, dominated by a coherent CSF-centred redox axis alongside multiple partially opposing peripheral components. Age stratification revealed progressive fragmentation: middle-aged adults retained four coherent cross-compartment factors, whereas older adults exhibited five more dispersed components. Sex-stratified analyses showed that females exhibited four-factor modular organisation centred on glutathione, while males showed a simpler three-factor structure with tighter cross-compartment coupling anchored by FRAP. Blood and CSF oxidative stress biomarkers are not interchangeable, a finding with direct implications for biomarker selection in clinical trials targeting neurological conditions.

**Highlights:** ⍰ Blood and CSF oxidative stress biomarkers show compartment-specific profiles

⍰ FRAP is the only marker significantly correlated across blood and CSF

⍰ Redox factor structure becomes more fragmented with aging

⍰ Sex-stratified analyses reveal divergent peripheral antioxidant organisation

⍰ Peripheral oxidative stress markers may not reflect CNS redox status

---

Aging involves the progressive buildup of cellular by-products and damage to proteins, DNA, and other cellular components, occurring at varying rates^1,2,3^. Among these is oxidative stress, which arises when the generation of prooxidant free radicals outpaces the system’s antioxidant defenses, particularly when prooxidant levels exceed those of antioxidants^1,4^. Oxidative stress levels tend to rise with age as antioxidant defenses decline, especially during the midlife transition^5,6,7^. Interestingly, the onset of chronic and neurodegenerative diseases in late midlife or older usually correlates with pathophysiological changes that begin during midlife^8,9,10^. This link arises because prolonged oxidative stress drives progressive injury to macromolecules and intracellular structures, gradually impairing key biological systems such as mitochondrial function, inflammatory regulation, and neuronal integrity, which, over time, predispose individuals to disease^11,12,13^. As a result, oxidative stress has been implicated in the pathogenesis of aging and contributes to a broad spectrum of chronic and degenerative conditions, including autoimmune disorders^14,15^, inflammation, cancer^16^, arthritis^17^, cardiovascular disease^18^, and neurodegenerative diseases^19^, making it a target for pharmacological intervention^20,21^. However, while preclinical results have shown promise^18^, most clinical trials targeting oxidative stress have failed to yield positive results in human studies^22^. This translational gap may stem in part from the overreliance on peripheral biomarkers, which do not always reflect redox dynamics within the central nervous system (CNS), where many age-related pathologies originate^16,23,24^. A crucial next step is to clarify the behavior of oxidative stress biomarkers in different populations, including across age groups^25^, sexes^26^, and chronic disease stages^27,28^. Most antioxidant studies to date have focused almost exclusively on blood-based markers, leaving CNS-specific redox processes underexplored^29,30^. While studies have shown inconsistent correspondence between blood and CSF biomarkers in other research areas, little is known about whether such discrepancies also apply to oxidative stress^31^.

Oxidative stress can be measured directly through reactive oxygen species (ROS) or indirectly through indices of lipid peroxidation, protein nitration/oxidation, and DNA/RNA damage^32,33^. Indirect biomarkers have proven to be more stable for long-term measurement^34^, but even among these, congruence across fluid modalities remains unclear. Understanding the physiological relationship between indirect oxidative stress biomarkers in blood and CSF could inform the design of more accurate diagnostic tools and improved management of age-related chronic diseases. Historically, research has focused on measuring oxidative stress through blood samples, while studies on oxidative stress in CSF have been limited, often focusing on protein damage from oxidative stress^35, 36, 37^. There is a growing need to more thoroughly characterize the correspondence between oxidative stress biomarkers across blood and CSF. This study examines the cross-compartment profile of oxidative stress biomarkers in blood and CSF across age and sex. We measured Total Antioxidant Capacity (TAC), γ-glutamylcysteinylglycine (Glutathione, GSH), Hydroxyl Radical Scavenging Activity (HRSA), and Ferric Reducing Antioxidant Power (FRAP), which reflect antioxidant capacity (higher values indicate greater antioxidant defense and lower oxidative stress), as well as thiobarbituric acid-reactive substances (TBARS), an index of lipid peroxidation (higher values reflect greater oxidative burden) (Table 1). We seek to understand the dynamics between these biomarkers across fluids and explore their interrelationships in blood and CSF. An improved understanding of these connections could support the development of more effective therapeutic and preventive approaches to oxidative stress-related and its associated chronic diseases.

**Table 1.**

| Biomarker | Units | Metabolic pathway | Biological Function | Interpretation | Modality |
| --- | --- | --- | --- | --- | --- |
| FRAP | mmol Fe<sup>2+</sup>/L | Redox Balance | Measures reducing power by reducing Fe<sup>3+</sup> to Fe<sup>2+</sup> | Higher means greater overall antioxidant capacity | Blood and CSF |
| GSH | μmol/L | Glutathione metabolism / Redox homeostasis | Major intracellular antioxidant, detoxifies ROS | Higher means better cellular antioxidant defense | Blood and CSF |
| TAC | mmol Trolox equivalent/L | Overall oxidative stress response | Measures the cumulative ability of antioxidants to neutralize free radicals | Higher means greater total antioxidant defense | Blood and CSF |
| TBARS | nmol MDA/mL (Malondialdehyde equivalent) | Lipid peroxidation | Measures oxidative damage to lipids | Higher means more lipid oxidation (indicative of oxidative damage) | Blood and CSF |
| HRSA | % Inhibition or μmol Trolox equivalent/L | ROS neutralization | Assesses the ability to scavenge hydroxyl radicals, preventing oxidative stress | Higher means better ability to neutralize hydroxyl radicals | Blood |

ROS: Reactive oxygen species. CSF: Cerebrospinal Fluid. TAC: Total Antioxidant Capacity, GSH: γ-glutamylcysteinylglycine or Glutathione, HRSA: Hydroxyl Radical Scavenging Activity, FRAP: Ferric Reducing Antioxidant Power, and TBARS: thiobarbituric acid-reactive substances.

## RESULTS

We collected biological samples of 140 participants who were stratified by age into middle-aged (<65 years) and older adults (≥65 years; Table 2A), and by sex (Table 2B). Older adults had significantly fewer years of education (*p* < .003) but were similar to middle-aged participants in sex distribution and in all blood and CSF oxidative stress biomarkers. Comparisons by sex revealed that males had significantly higher blood levels of total antioxidant capacity (TAC; *p* = .004) and glutathione (GSH; *p* = .017) than females. No other blood biomarkers or any CSF oxidative stress markers differed significantly between sexes. Overall, redox biomarker profiles were largely consistent across age and sex, with the exception of higher peripheral antioxidant levels observed in men.

**Table 2A.** Baseline characteristics of participants in the analysis of blood vs cerebrospinal fluid congruence among age groups.

|  | Total population (n=140) | Middle-aged (n=68) | Older adults (n=72) | *p-value* |
| --- | --- | --- | --- | --- |
| **Demographics<sup>a</sup>** |  |  |  |  |
| Age (years), mean (SD) | 64.02 (9.21) | 56.22 (5.90) | 71.28 (4.67) | <.001* |
| Male Sex, n (%) | 47 (33.60) | 21 (30.90) | 26 (36.10) | .437 |
| Education (years), mean (SD) | 13.2 (3.96) | 14.22 (3.53) | 12.25 (4.11) | <.003* |
| **Blood redox biomarker levels<sup>b</sup>** |  |  |  |  |
| TAC (mmol Trolox equivalent/L) | .96 (.11) | .96 (.12) | .96 (.09) | .752 |
| GSH (μmol/L) | 11.81 (4.33) | 12.27 (4.65) | 11.39 (3.99) | .380 |
| TBARS (nmol MDA/mL (Malondialdehyde equivalent)) | 5.01 (1.68) | 5.02 (1.66) | 5.00 (1.72) | .797 |
| FRAP (mmol Fe<sup>2+</sup>/L) | .43 (.09) | .42 (.10) | .44 (.08) | .170 |
| HRSA (% Inhibition or μmol Trolox equivalent/L) | .02 (.00) | .02 (.00) | .02 (.00) | .781 |
| **Cerebrospinal fluid redox biomarker levels<sup>b</sup>** |  |  |  |  |
| TAC (mmol Trolox equivalent/L) | .37 (.135) | .36 (.14) | .38 (.13) | .401 |
| GSH (μmol/L) | 13.3 (5.5) | 12.85 (5.21) | 13.74 (5.77) | .361 |
| TBARS (nmol MDA/mL (Malondialdehyde equivalent)) | 1.51 (1.00) | 1.46 (.97) | 1.56 (1.04) | .696 |
| FRAP (mmol Fe<sup>2+</sup>/L) | .29 (.07) | .29 (.06) | .30 (.07) | .527 |

TAC: Total Antioxidant Capacity, GSH: γ-glutamylcysteinylglycine or Glutathione, HRSA: Hydroxyl Radical Scavenging Activity, FRAP: Ferric Reducing Antioxidant Power, and TBARS: thiobarbituric acid-reactive substances.
<sup>a</sup>T-test, <sup>b</sup>Mann-U Whitney.

**Table 2B.** Baseline characteristics of participants in the analysis of blood vs cerebrospinal fluid congruence among sexes.

|  | Total population (n=140) | Female (n=93) | Male (n=47) | *p-value* |
| --- | --- | --- | --- | --- |
| **Demographics<sup>a</sup>** |  |  |  |  |
| Age (years), mean (SD) | 64.02 (9.21) | 63.47 (9.15) | 65.06 (9.26) | .325 |
| Education (years), mean (SD) | 13.2 (3.96) | 13.26 (3.76) | 13.09 (4.37) | .821 |
| **Blood oxidative stress biomarker levels<sup>b</sup>** |  |  |  |  |
| TAC (mmol Trolox equivalent/L) | .96 (.11) | .944 (.10) | 1.00 (.11) | .004* |
| GSH (μmol/L) | 11.81 (4.33) | 11.18 (4.14) | 13.09 (4.46) | .017* |
| TBARS (nmol MDA/mL (Malondialdehyde equivalent)) | 5.01 (1.68) | 5.10 (1.63) | 4.83 (1.79) | .315 |
| FRAP (mmol Fe<sup>2+</sup>/L) | .43 (.09) | .43 (.09) | .43 (.09) | .788 |
| HRSA (% Inhibition or μmol Trolox equivalent/L) | .02 (.00) | .018 (.00) | .02 (.00) | .137 |
| **Cerebrospinal fluid oxidative stress biomarker levels<sup>b</sup>** |  |  |  |  |
| TAC (mmol Trolox equivalent/L) | .37 (.14) | .36 (.13) | .38 (.15) | .603 |
| GSH (μmol/L) | 13.3 (5.50) | 13.32 (5.70) | 13.30 (5.15) | .874 |
| TBARS (nmol MDA/mL<br>(Malondialdehyde equivalent)) | 1.51 (1.00) | 1.54 (1.05) | 1.46 (.92) | .797 |
| FRAP (mmol Fe<sup>2+</sup>/L) | .29 (.07) | .29 (.07) | .29 (0.07) | .962 |

Total Antioxidant Capacity (TAC), γ-glutamylcysteinylglycine (Glutathione, GSH), Hydroxyl Radical Scavenging Activity (HRSA), Ferric Reducing Antioxidant Power (FRAP), and thiobarbituric acid-reactive substances (TBARS). <sup>a</sup>T-test, <sup>b</sup>Mann-U Whitney.

### Blood-CSF Biomarker Relationships

To assess intercompartmental dynamics of oxidative stress, we examined Spearman correlations between blood and CSF concentrations of homologous biomarkers (Figure 2). Among the four biomarker pairs, only Ferric Reducing Antioxidant Power (FRAP-b vs. FRAP-csf) showed a significant positive association (ρ = +0.49, FDR-p < 0.001), indicating that blood-based FRAP partially reflects central nervous system antioxidant capacity.

**Fig. 1.**
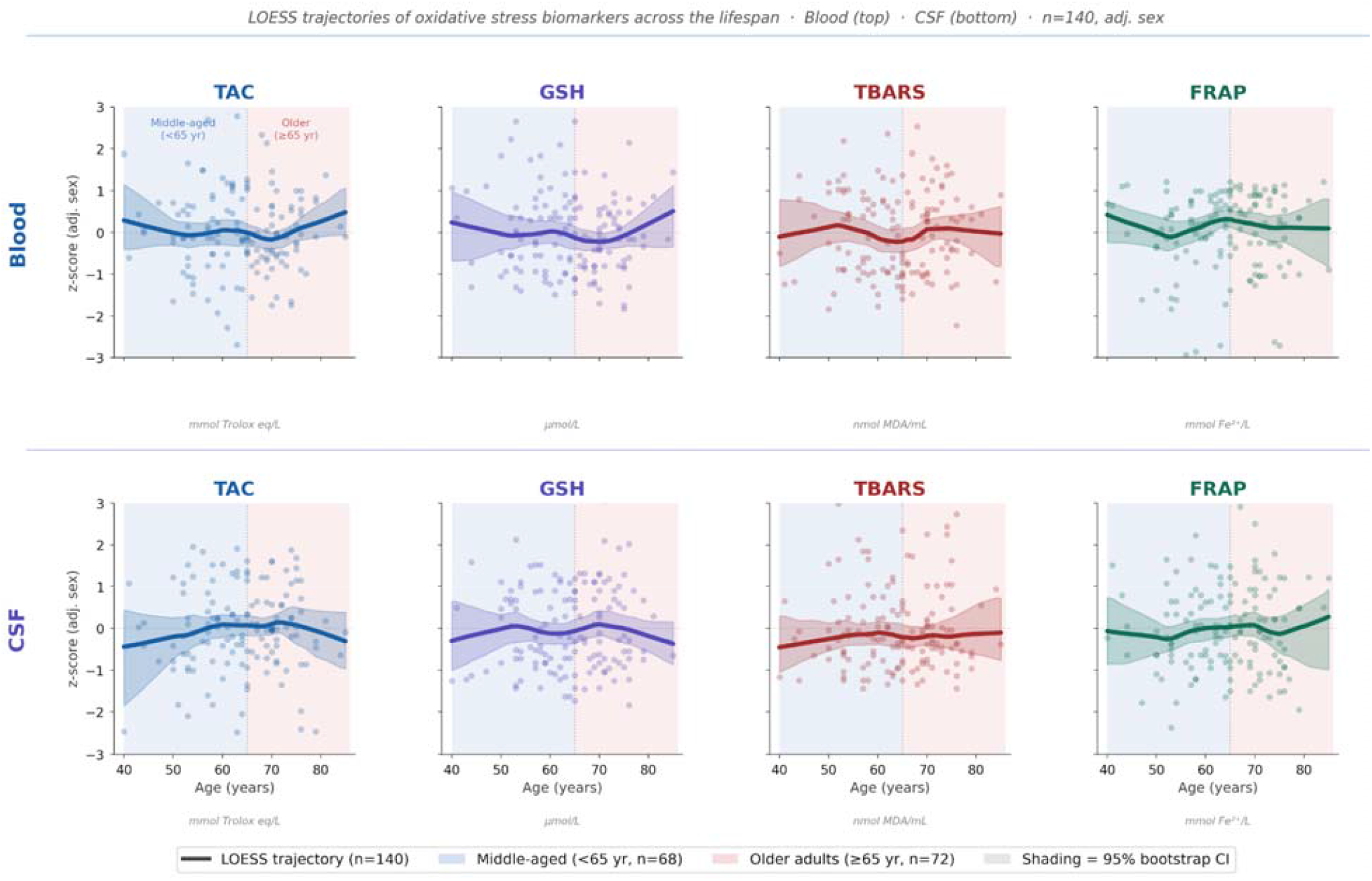
LOESS trajectories of oxidative stress biomarkers across the lifespan in blood and cerebrospinal fluid. Locally estimated scatterplot smoothing (LOESS) curves depict the age-related trajectories of four oxidative stress biomarkers measured in blood (top row) and cerebrospinal fluid (CSF; bottom row) across the lifespan (n=140, age range 40–85 years). Individual observations are shown as semi-transparent dots. The solid line represents the LOESS trajectory (bandwidth = 0.55) and the shaded band represents the 95% bootstrap confidence interval (150 iterations). Biomarker values were winsorised at ±3 SD, z-score standardised, and adjusted for sex by regression prior to smoothing. Background shading denotes age strata: middle-aged adults (<65 years, n=68, blue) and older adults (≥65 years, n=72, red); the dotted vertical line marks the stratum boundary at age 65. Biomarkers are displayed in matched column order across compartments to facilitate visual comparison: Total Antioxidant Capacity (TAC; mmol Trolox eq/L), Glutathione (GSH; μmol/L), Thiobarbituric Acid-Reactive Substances (TBARS; nmol MDA/mL), and Ferric Reducing Antioxidant Power (FRAP; mmol Fe^2+^/L). Hydroxyl Radical Scavenging Activity (HRSA) was measured in blood only and is omitted from this figure given near-zero variance across the sample. The largely flat trajectories across both compartments indicate that individual biomarker levels do not show strong monotonic age trends, motivating the latent variable analyses presented in subsequent figures.

**Fig 2.**
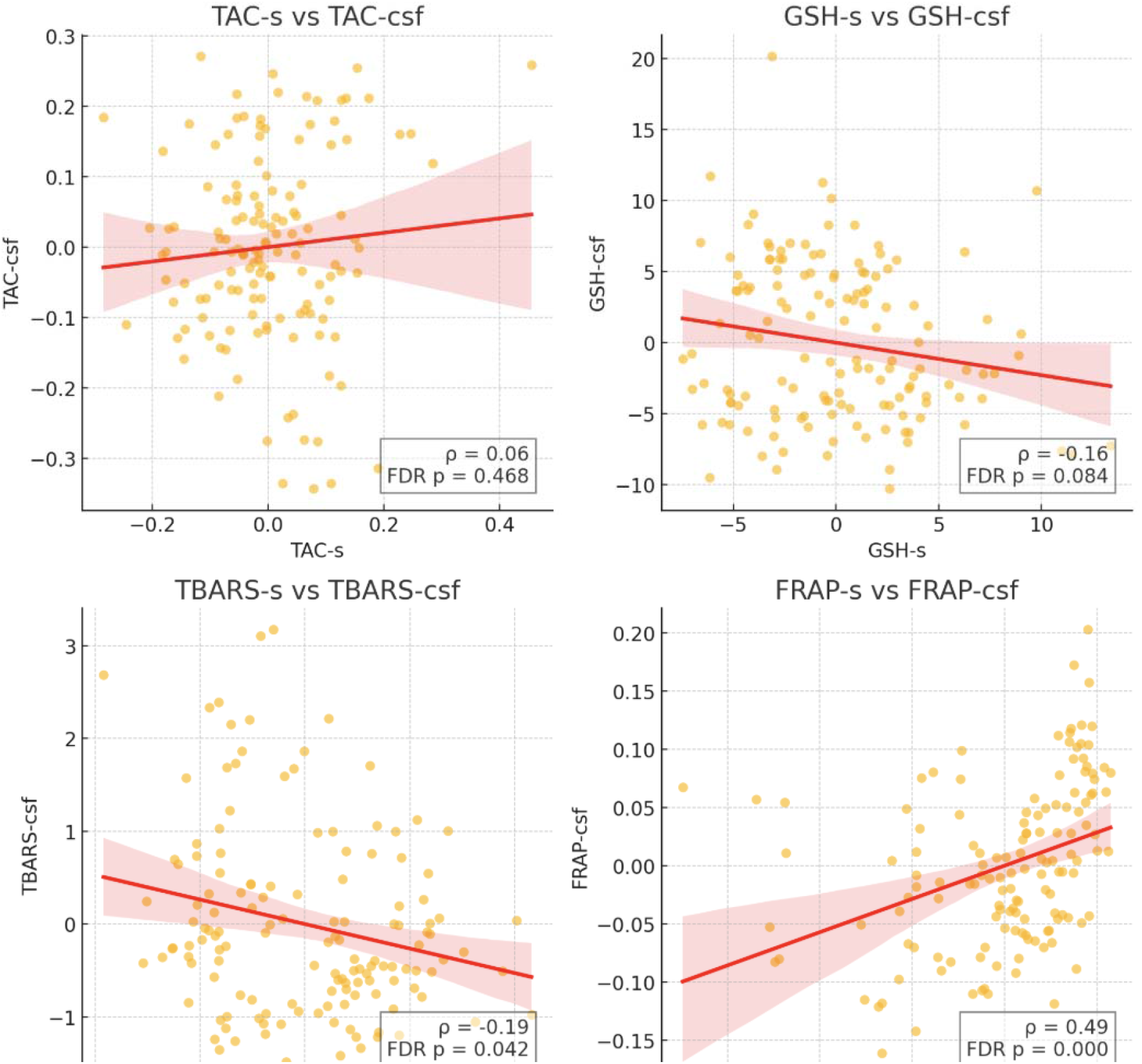
Intercompartmental relationship between oxidative stress biomarkers. Scatterplots show adjusted Spearman correlations between blood (x-axis) and cerebrospinal fluid (CSF; y-axis) levels of key oxidative stress biomarkers. All correlations are adjusted for age and sex. Shaded areas represent 95% confidence intervals. **a**. Total Antioxidant Capacity (TAC-b vs. TAC-csf), b. Glutathione (GSH-b vs. GSH-csf), **c**. Lipid peroxidation (TBARS-b vs. TBARS-csf), **d**. Reducing power (FRAP-b vs. FRAP-csf). Spearman’s rho (ρ) and FDR-adjusted p-values are reported.

A modest inverse correlation was observed for thiobarbituric acid-reactive substances (TBARS-b vs. TBARS-csf), potentially reflecting a compensatory mechanism in lipid peroxidation regulation between compartments. The relationship between glutathione concentrations (GSH-b vs. GSH-csf) was weak and non-significant (ρ = −0.16, FDR-p = 0.158), and Total Antioxidant Capacity (TAC-b vs. TAC-csf) showed minimal intercompartmental alignment (ρ = +0.06, FDR-p = 0.520).

Collectively, these findings highlight that while blood-based FRAP may be subject to systemic regulatory mechanisms, other processes-particularly glutathione metabolism and lipid peroxidation may exhibit limited peripheral-central congruence, underscoring the complexity of oxidative stress regulation pathways across biological fluids.

### Latent Structures of Oxidative Stress Biomarkers in Blood and CSF

Oxidative stress results from the interplay of multiple biological systems working in concert across tissues. While individual correlations between biomarkers (e.g., GSH or FRAP) help identify pairwise associations, they do not capture the broader, coordinated patterns of regulation that may underlie systemic or compartmentalized oxidative stress responses.

To uncover these deeper patterns, we used factor analysis, a statistical method that groups correlated variables into underlying dimensions called latent factors. Each factor represents a shared biological mechanism that explains the behavior of several biomarkers. A biomarker’s “loading” on a factor indicates how strongly it reflects that latent process. This approach allows us to identify clusters of oxidative stress biomarkers that co-vary, potentially reflecting shared metabolic pathways or compartment-specific redox regulation. It also helps clarify whether blood-based markers reflect central oxidative dynamics or operate independently.

We conducted a factor analysis of blood and CSF biomarkers using principal component extraction with varimax rotation (Figure 3). Four factors with eigenvalues >1 were retained, capturing a total of 66.6% of the variance in oxidative stress profiles. The proportion of variance explained by each factor is shown below the corresponding factor in Figure 3. Only biomarkers with factor loadings greater than ±0.4 were considered to meaningfully contribute to each factor.

**Fig 3.**
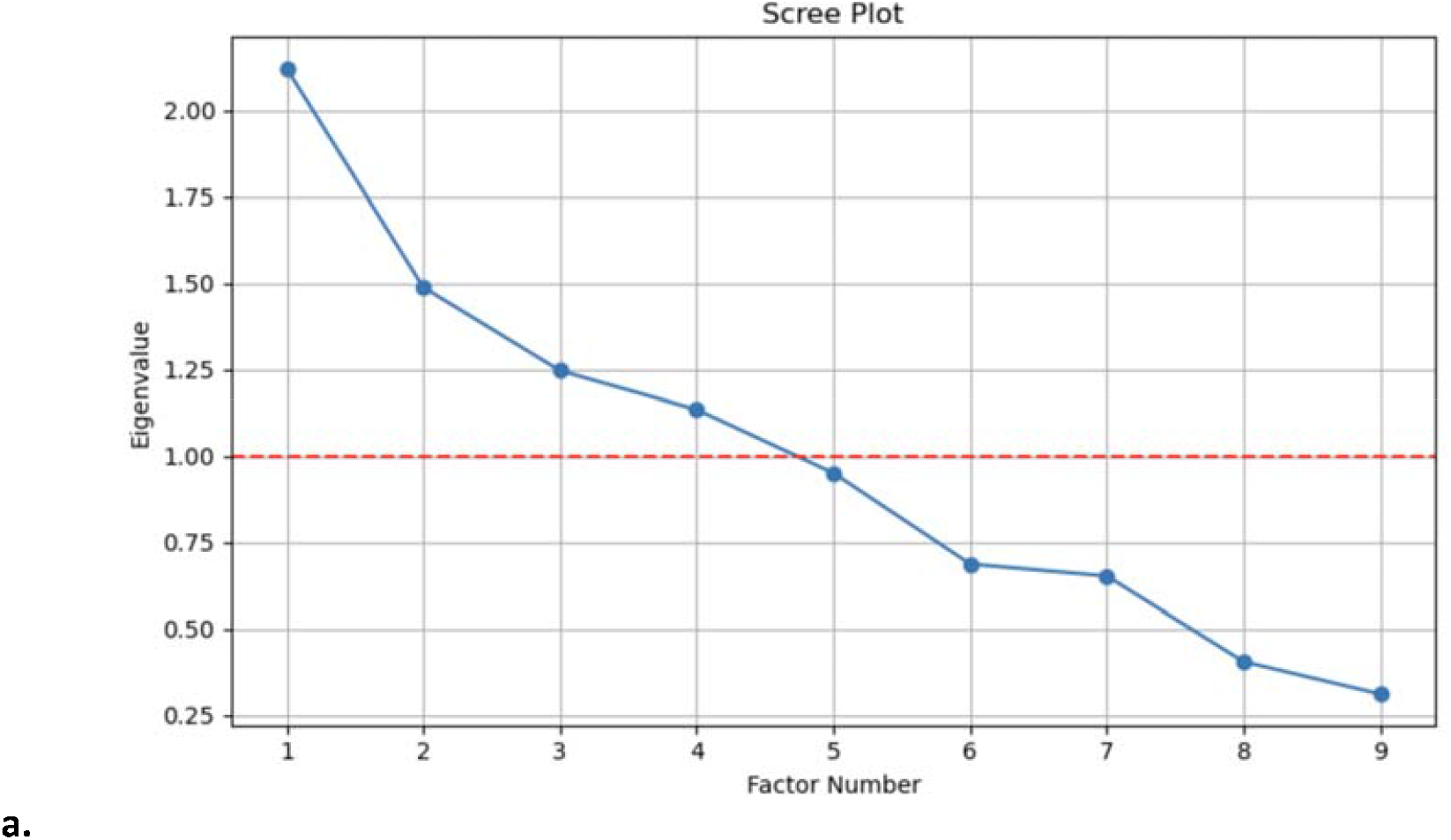
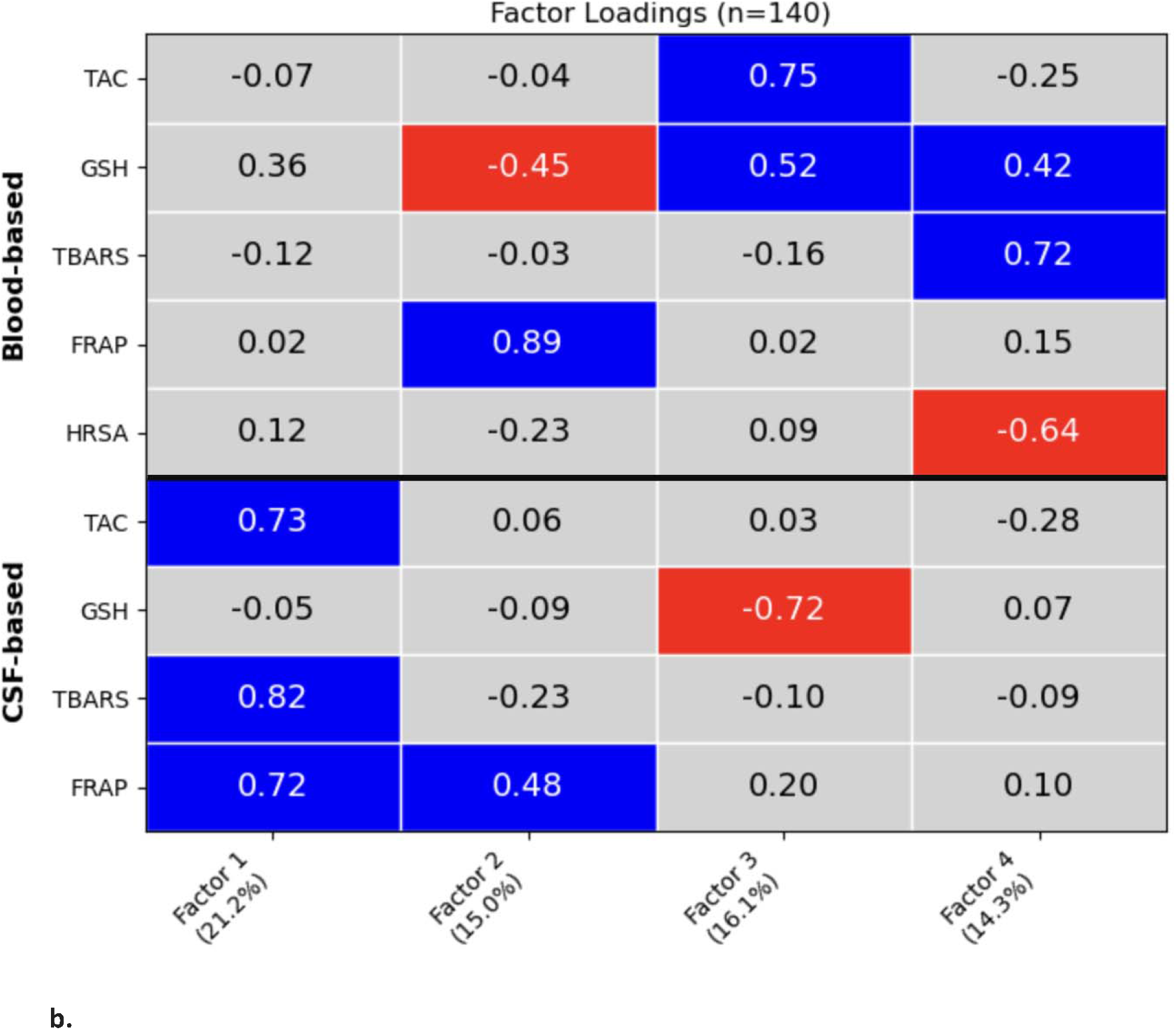
Factor analysis for Oxidative Stress biomarkers in Blood and CSF. **a**. Scree Plot. Displays the eigenvalues for each extracted factor. Four factors were retained (eigenvalues > 1), indicating they explain meaningful variance in biomarker patterns. **b**. Factor Loadings (Varimax Rotated), with factor loading magnitude interpreted using a ±0.4 threshold. Heatmap shows how nine biomarkers load onto the four factors. High loadings (≥ ±0.4) suggest meaningful contributions to latent oxidative stress mechanisms across fluids. Blue shades reflect positive meaningful contributions; red shades indicate negative meaningful contributions; intermediate values are **gray**. Each factor label includes the percentage of variance it explains in parentheses. Biomarkers include blood and CSF measures of TAC (Total Antioxidant Capacity), GSH (Glutathione), TBARS (Lipid peroxidation), FRAP (Reducing power), HRSA (Hydroxyl radical scavenging activity). **All biomarker values were adjusted for age and sex prior to factor analysis.**

Factor 1 was driven predominantly by CSF biomarkers (TAC, TBARS, and FRAP) and likely reflects a CSF-centered oxidative stress axis, integrating both antioxidant defenses and lipid peroxidation in the central nervous system.

Factor 2 was defined by strong positive loadings from FRAP-b (+0.89) and FRAP-csf (+0.48), and a negative loading from GSH-b (−0.45). This cross-compartmental FRAP axis may reflect a global antioxidant mechanism anchored in ferric reducing capacity, while the inverse association with peripheral GSH suggests potentially divergent or compensatory regulation between thiol- and non-thiol-based antioxidant pathways.

Factor 3 included positive loadings from TAC-b and GSH-b, and a negative loading from GSH-csf. This suggests a coordinated upregulation of peripheral antioxidant capacity (via total antioxidant capacity and glutathione) alongside a suppression of central glutathione activity, highlighting compartment-specific glutathione dynamics.

Factor 4 was characterized by positive contributions from GSH-b and TBARS-b, alongside a negative loading from HRSA-b. This pattern may reflect a complex interplay of oxidative damage and antioxidant compensation in the periphery, where increased lipid peroxidation and glutathione are paired with reduced hydroxyl radical scavenging activity.

Collectively, these findings underscore that while a unified CSF oxidative stress axis exists, peripheral oxidative markers are distributed across multiple, partially opposing latent factors. The joint appearance of FRAP-b and FRAP-csf in a single factor supports its role as a potential cross-compartment oxidative stress marker, in contrast to glutathione, which demonstrates opposing regulation between compartments.

### Latent Structures of Oxidative Stress Biomarkers by Age

To examine age-related differences in oxidative stress regulation, we conducted separate factor analyses for middle-aged and older adults, adjusted for sex (Figure 4).

**Fig 4.**
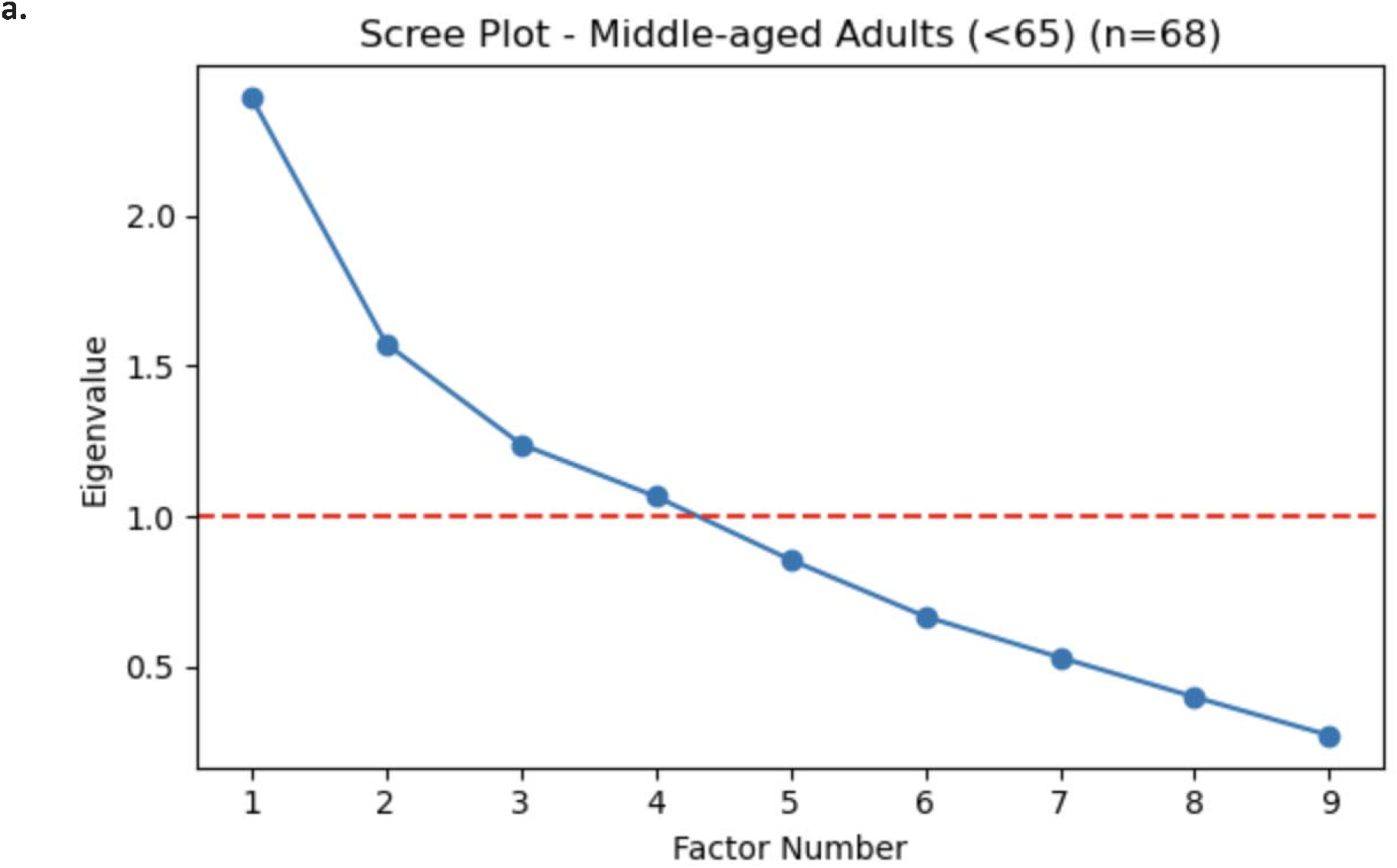
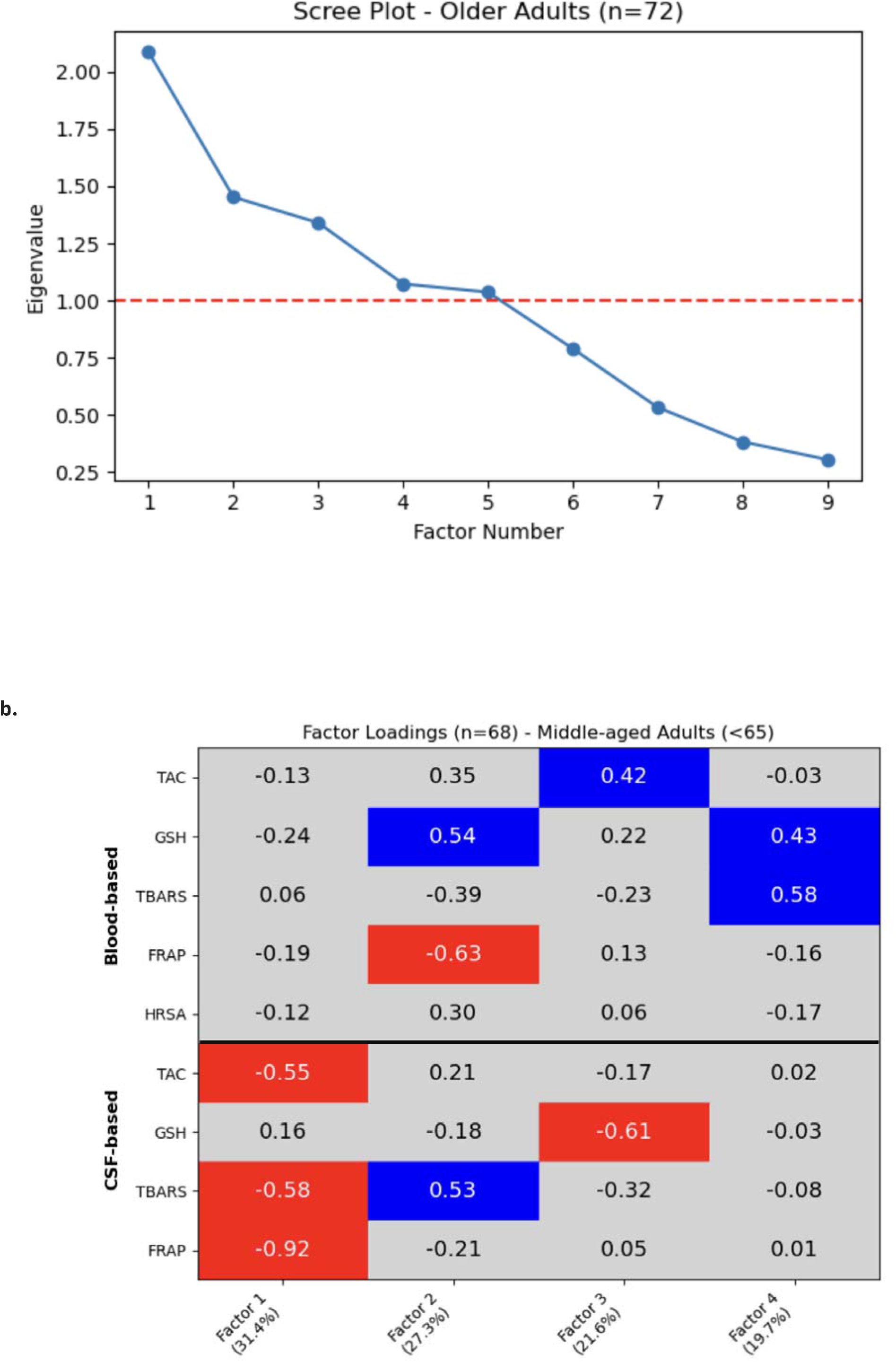
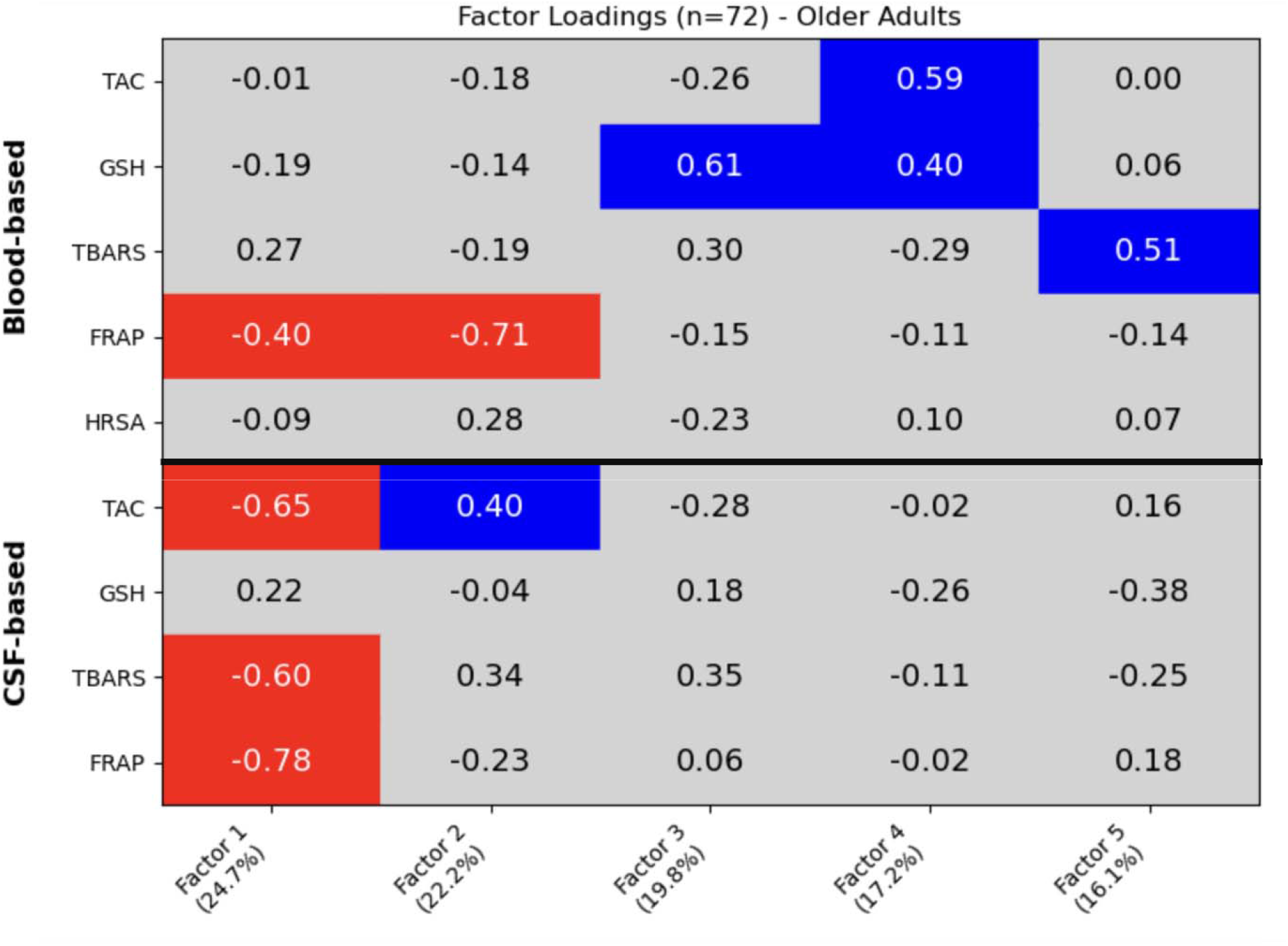
Factor analysis per age group. **a**. Scree plots show the number of latent factors (eigenvalues >1) retained using maximum likelihood factor extraction without rotation. **b**. Heatmaps display factor loadings for oxidative stress biomarkers in middle-aged adults (< 65 years, n = 68) and older adults (≥ 65 years, n = 72), with factor loading magnitude interpreted using a ±0.4 threshold. **Blue** shades reflect positive meaningful contributions; **red** shades indicate negative meaningful contributions; intermediate values are **gray**. Biomarkers include: TAC-b = Total Antioxidant Capacity, GSH-b = Glutathione, TBARS-b = Lipid peroxidation, FRAP-b = Ferric Reducing Antioxidant Power, HRSA-b = Hydroxyl Radical Scavenging Activity, -b = In Blood. TAC-csf, GSH-csf, TBARS-csf, FRAP-csf = respective markers in cerebrospinal fluid (CSF). **All biomarker values were adjusted for sex prior to factor analysis**.

In middle-aged adults, several factors closely resembled the latent structure observed in the general population sample. Factor 3 preserved the co-loading of TAC-b and GSH-b, alongside a negative loading from GSH-csf, replicating the peripheral antioxidant axis coupled with central glutathione opposition seen in the full group. Similarly, Factor 2 retained a prominent negative loading from FRAP-b, consistent with its isolated peripheral antioxidant role. Although FRAP-csf did not contribute meaningfully to this factor as it did in the general population, its absence may reflect the early onset of cross-compartmental decoupling.

Factor 1 in middle-aged adults displayed opposite loading direction compared to the general population sample, with TAC-csf, TBARS-csf, and FRAP-csf all loading negatively. However, the presence of these same three CSF biomarkers within the factor suggests preserved structural coherence. This indicates that while the composition of central oxidative markers remains intact, their relationship to the underlying latent factor may shift in midlife, potentially due to changes in regulatory dynamics or early dampening of central antioxidant defenses.

Factor 4 diverged more markedly, combining TBARS-b, GSH-b, and GSH-csf, with the latter again loading negatively. This pattern indicates the emergence of a lipid peroxidation–glutathione axis spanning both compartments, potentially signaling the beginning of compensatory shifts in redox balance.

In older adults, the latent structure became more fragmented, with five factors retained and broader dispersion of biomarker loadings. While Factor 2 preserved positive loadings for TAC-csf and GSH-csf, and Factor 3 retained the negative GSH-b loading seen in younger participants, the remaining components exhibited greater divergence. Factor 1 clustered strong negative CSF-based loadings from TAC, TBARS, and FRAP, suggesting unified downregulation of central antioxidant capacity with age. FRAP-b loaded negatively across multiple factors (1 and 2), indicating a breakdown in its prior cross-compartment association. Factors 4 and 5 showed weaker structure but featured residual peripheral processes; Factor 4 via modest TAC-b loading, and Factor 5 via isolated TBARS-csf contribution.

Together, these findings suggest that while middle-aged adults maintain a more integrated oxidative stress architecture, similar to that of the general population sample, older adults exhibit a more fragmented and compartmentalized latent structure. Biomarkers that previously co-loaded, particularly those related to glutathione and total antioxidant capacity, become increasingly dissociated with age. This shift may reflect diminished inter-compartmental redox coordination or compensatory adaptations in response to accumulating oxidative stress over the lifespan.

### Sex-Specific Latent Structures of Oxidative Stress Biomarkers

To explore whether oxidative stress regulation differs by sex, we conducted age-adjusted factor analyses separately in males and females (Figure 5), comparing the resulting latent structures to those observed in the total sample (n=140). This approach enabled the identification of shared components, sex-specific divergences, and newly emergent patterns within each subgroup.

**Fig 5.**
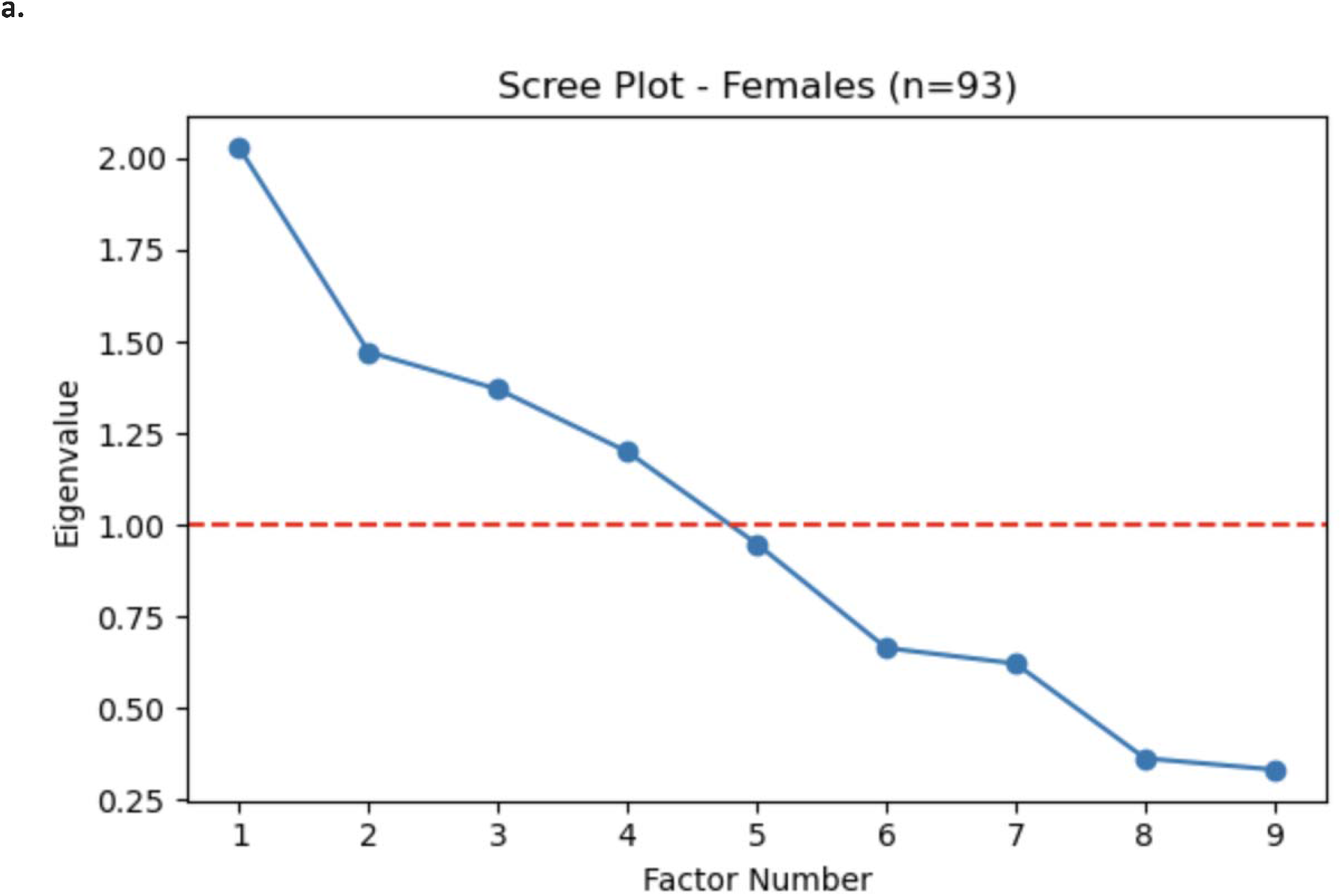
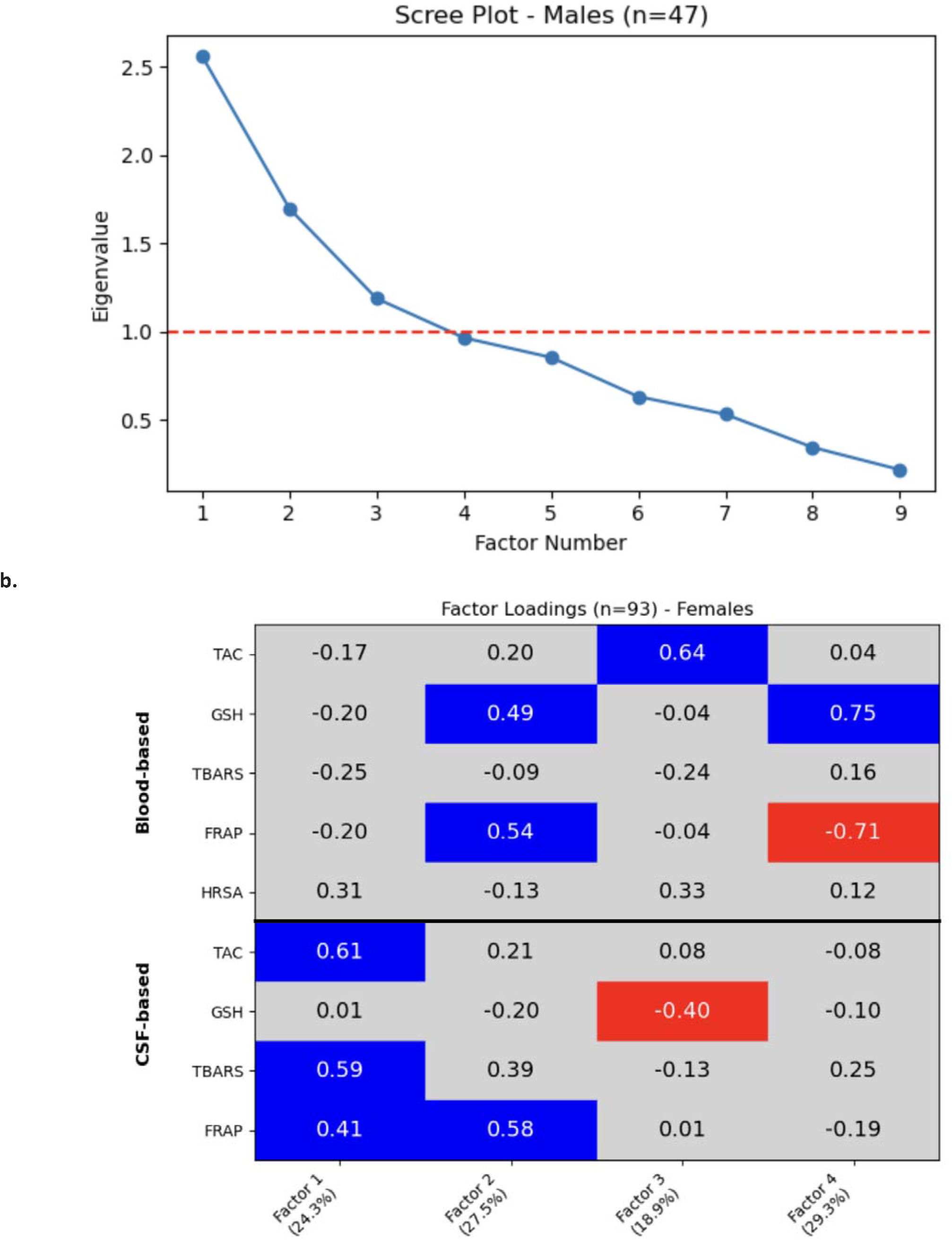
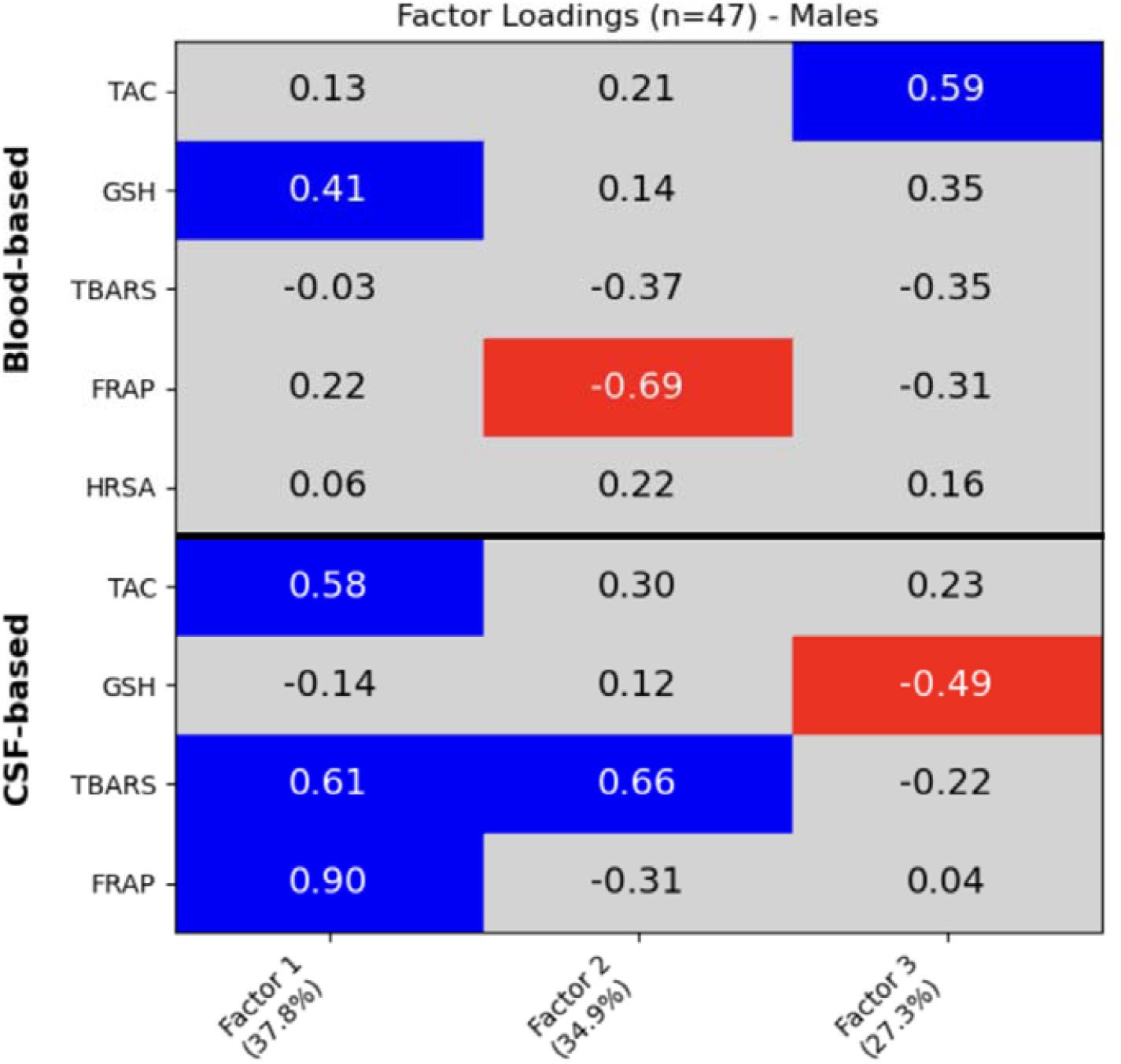
Factor analysis per sex group. **a**. Scree plots show the eigenvalues from maximum likelihood factor extraction in females (n = 93) and males (n = 47). Factors were retained using the Kaiser criterion (eigenvalue >1). For males, three factors were retained; parallel analysis supported two factors empirically, and the third factor (EV = 1.19) was retained on theoretical grounds. No rotation was applied. **b**. Heatmaps display rotated factor loadings of oxidative stress biomarkers, with factor loading magnitude interpreted using a ±0.4 threshold. Analyses were adjusted for age. **Blue** shades reflect positive meaningful contributions; **red** shades indicate negative meaningful contributions; intermediate values are **gray**. Biomarkers include: TAC-b = Total Antioxidant Capacity, GSH-b = Glutathione, TBARS-b = Lipid peroxidation, FRAP-b = Ferric Reducing Antioxidant Power, HRSA-b = Hydroxyl Radical Scavenging Activity, -b = In Blood. TAC-csf, GSH-csf, TBARS-csf, FRAP-csf = respective markers in cerebrospinal fluid (CSF). **All biomarker values were adjusted for age prior to factor analysis**.

**Fig 6.**
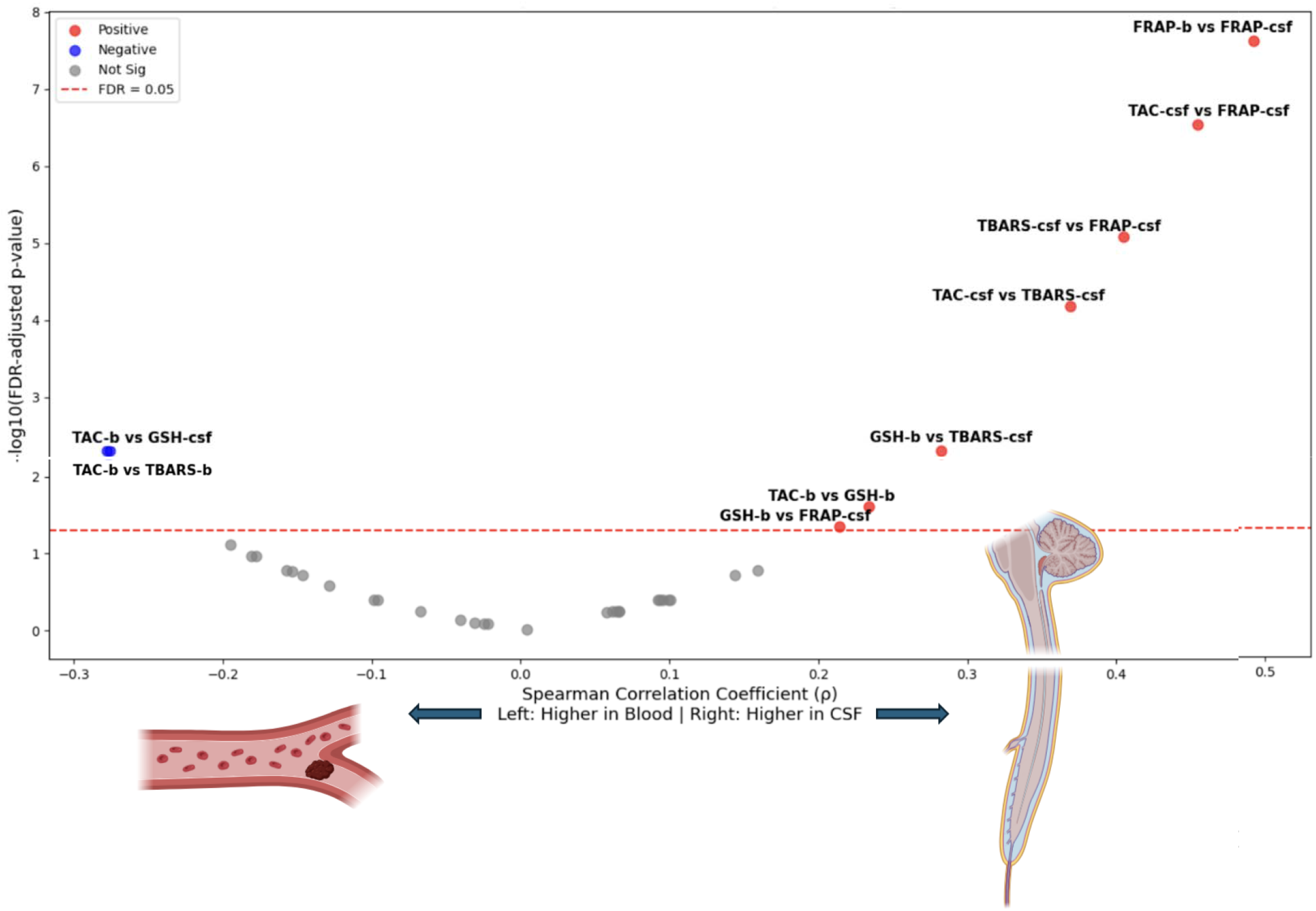
Systemic-Central Relationships Among Redox Biomarkers in Blood and CSF. Volcano plot of correlation coefficients (x axis, ρ) versus P value (y axis, −log10 scale) for the whole cohort, including all correlations for blood-based and CSF oxidative stress biomarkers. Red values indicate correlations where CSF and blood oxidative stress biomarkers were positively associated, Blue values indicate correlations where CSF and Blood were inversely associated. Grey values indicate non-significant correlations. The horizontal line represents the FDR-adjusted p-value at 0.05. Total Antioxidant Capacity (TAC), γ-glutamylcysteinylglycine (Glutathione, GSH), Hydroxyl Radical Scavenging Activity (HRSA), Ferric Reducing Antioxidant Power (FRAP), and thiobarbituric acid-reactive substances (TBARS).

In females (n=93), four factors were retained. Factor 1 replicated the CSF-centered oxidative stress axis. Factor 2 also closely mirrored the general structure, indicating a preserved antioxidant-peroxidation coupling across compartments. These consistencies suggest stability in central redox architecture in females. However, Factor 3 diverged meaningfully: it was uniquely characterized by a negative loading of GSH-csf, with no strong blood biomarker contributors. This contrasts with the general sample, where GSH-csf opposed a peripheral antioxidant axis involving GSH-b and TAC-b. Its isolation in females may indicate a decoupling of central glutathione dynamics or compensatory regulation in the CSF compartment. Factor 4, in contrast to the general population structure, did not include TAC-b, but retained a strong positive loading from GSH-b and a negative loading from FRAP-b, suggesting a competitive or opposing interaction between peripheral glutathione activity and reducing capacity in females. Notably, GSH-b maintained a distinct presence in multiple factors, reinforcing its central role in systemic redox regulation in this subgroup.

In males (n=47), three factors were retained based on the Kaiser criterion (eigenvalues: F1=2.56, F2=1.70, F3=1.19). Factor 1 preserved the hallmark CSF-driven oxidative stress axis (FRAP-csf +0.90, TBARS-csf +0.61, TAC-csf +0.58), and additionally included a positive loading from GSH-b (+0.41), indicating that in males, peripheral glutathione activity is partially integrated into the central antioxidant-peroxidation architecture, a pattern more cross-compartmental than observed in females or the full sample. Factor 2 reflected an opposition between peripheral reducing capacity and central lipid peroxidation (FRAP-b −0.69, TBARS-csf +0.66), suggesting that suppression of systemic reducing activity may co-occur with elevated central oxidative damage. Factor 3 captured an opposing gradient between peripheral antioxidant capacity and central glutathione (TAC-b +0.59, GSH-csf −0.49), pointing to possible compartmental decoupling of redox control whereby enhanced systemic antioxidant reserves are accompanied by reduced central glutathione availability.

In summary, both shared and sex-specific patterns emerged in the latent structure of oxidative stress biomarkers. While the CSF-centered antioxidant-peroxidation axis was preserved across sexes, peripheral redox organization diverged: females showed more modular glutathione dynamics, whereas males exhibited more integrative but opposing cross-compartment profiles. These findings highlight the value of sex-stratified modeling and may reflect underlying hormonal or neuroimmune differences.

### Cross-Compartment Correlations Among Oxidative Stress Biomarkers

To examine the degree of coordination between systemic and central oxidative stress processes, we conducted pairwise Spearman correlations between all blood- and CSF-based biomarkers, adjusting for age and sex and correcting for multiple comparisons using the false discovery rate (FDR; Fig. 5). Several significant cross-compartment associations emerged, with patterns that align closely with the latent factors identified in the full sample (Fig. 2).

FRAP-b and FRAP-csf showed the strongest positive correlation (ρ = +0.49, FDR-p < 0.001), consistent with their shared contribution to Factor 3, which captured a cross-compartment antioxidant dimension centered on ferric reducing activity. Similarly, TAC-csf was significantly correlated with both FRAP-csf and TBARS-csf, all of which clustered within Factor 1, the CSF-centered oxidative stress axis. These findings reinforce the coherence of central redox signaling and suggest that TAC-csf may bridge antioxidant and peroxidation processes in the central nervous system.

GSH-b showed a significant positive correlation with TBARS-csf (ρ = +0.28, FDR-p = 0.006), linking two biomarkers that otherwise loaded onto distinct factors-Factor 2 for GSH-b and Factor 1 for TBARS-csf-indicating a partial peripheral-central relationship between glutathione availability and lipid peroxidation. In contrast, a negative correlation was observed between TAC-b and GSH-csf (ρ = −0.28, FDR-p = 0.006), while GSH-b showed a positive trend with FRAP-csf (ρ = +0.21, FDR-p = 0.055). These inverse relationships mirror patterns in Factor 4, where TAC-b and GSH-csf exhibited diverging loadings, and may reflect regulatory opposition between peripheral antioxidant stores and central oxidative activity.

Overall, these findings highlight FRAP as a consistently coordinated antioxidant signal across blood and CSF, aligning with its prominent cross-compartment role in the factor structure. In contrast, GSH and TAC appear more compartment-specific, supporting the notion that distinct fluid environments modulate oxidative stress regulation differently.

## Discussion

### Intercompartmental Relationships and Biomarker Congruence

The present findings advance our understanding of compartmental dynamics in oxidative stress, highlighting the distinct yet intersecting roles of blood and CSF biomarkers. The moderate cross-compartment correlation observed for FRAP highlights its potential as a shared indicator of antioxidant capacity across systemic and central compartments^38^. This finding aligns with prior work suggesting that FRAP captures generalized redox activity and may bridge peripheral and central oxidative stress profiles^39,40^.

In contrast, the weak correspondence between other homologous biomarkers-such as TAC and GSH-supports the idea that oxidative stress regulation is largely compartmentalized^41^. These markers likely reflect local metabolic demands^42^, tissue-specific redox environments^43^, and selective permeability of the blood-brain barrier (BBB)^44^. The discordance observed between TAC-b and TAC-csf, and between GSH-b and GSH-csf, suggests that peripheral measurements of these antioxidants may not reliably reflect central redox status-an important consideration for biomarker-guided diagnostics^45,46,47^.

Interestingly, the modest inverse correlation between TBARS-b and TBARS-csf may represent a compensatory mechanism, whereby peripheral lipid peroxidation is buffered by central antioxidant responses^48,49^. This is consistent with proposed models of redox balancing across compartments in the face of localized oxidative stress^34,41^.

### Aging and Oxidative Stress Coordination

Age-stratified analyses showed that the coordination between oxidative stress markers becomes less organized with aging^50,51^. In middle-aged adults, the structure of oxidative stress biomarkers closely resembled that of the full cohort^52,53^, with biomarkers grouping into clear, coherent patterns^54^. This suggests that younger individuals maintain a more integrated redox regulation system across blood and CSF.

In older adults, however, five distinct factors emerged, indicating a more fragmented architecture. While elements of the original structure persisted, including a CSF-centered oxidative stress axis and isolated glutathione dynamics. New patterns also emerged: one factor combined TBARS-csf with negatively loaded TAC-b, and another was driven primarily by TBARS-b, reflecting a distinct peripheral lipid peroxidation process not seen in younger individuals^55^. These changes align with known aging-related disruptions in redox balance, such as mitochondrial dysfunction^56^, blood-brain barrier breakdown^57^, and reduced antioxidant enzyme activity^55^.

Our findings suggest that with aging, redox coordination between compartments declines, possibly due to changes in BBB permeability or compartmental shifts in mitochondrial and antioxidant responses. This loss of integration may increase vulnerability to oxidative damage and contribute to age-related neurological decline.

### Sex Differences in Oxidative Stress Profiles

Our age⍰adjusted, sex⍰stratified factor analyses revealed both shared and divergent oxidative stress patterns across blood and CSF. While both sexes consistently retained a CSF⍰centered axis, driven by strong loadings of TAC⍰csf, TBARS⍰csf, and FRAP⍰csf, peripheral antioxidant structure diverged substantially.

In females, the peripheral redox architecture was notably modular. One latent factor was defined by positive loadings from TAC⍰b and a negative loading from GSH⍰csf, suggesting a compensatory upregulation of systemic antioxidant reserves in response to diminished central glutathione availability. Another factor paired strong positive contributions from GSH⍰b with a negative loading from FRAP⍰b, indicating potentially competing roles of thiol-based and non-thiol antioxidant systems. These patterns are consistent with experimental and clinical evidence that estrogens upregulate key redox-regulatory enzymes, including GPx, and promote glutathione synthesis and recycling^26, 58^. Animal models have shown that 17β-estradiol modulates redox-sensitive transcription factors like Nrf2, increasing systemic antioxidant defenses and reducing mitochondrial oxidative stress^59^.

In contrast, the male redox structure appeared more integrative yet functionally divergent. In one factor, peripheral GSH⍰b co-loaded with central antioxidant and peroxidation markers (FRAP⍰csf, TBARS⍰csf, TAC⍰csf), indicating tighter cross-compartment coupling. Another male-specific factor revealed an inverse gradient: TAC⍰b and TBARS⍰csf loaded positively, while TBARS⍰b and FRAP⍰b loaded negatively, suggesting that increased peripheral antioxidant capacity and central lipid peroxidation may emerge in parallel with suppression of other systemic defenses. These configurations may reflect testosterone’s dualistic role in oxidative regulation^60^. While testosterone has been shown to increase oxidative stress markers in various tissues, it may also upregulate antioxidant activity via androgen receptor signaling or peripheral conversion to estrogens, modulating Nrf2-dependent pathways under certain conditions^61^.

Together, these findings advance the concept of sex-specific redox architecture: females exhibit more modular and compartmentalized antioxidant strategies centred on glutathione across four factors, while males show a more parsimonious structure (three factors) with tighter cross-compartment integration anchored by FRAP and opposing peripheral-central gradients. These distinctions are biologically grounded in hormonal, metabolic, and mitochondrial regulatory differences and underscore the importance of sex-stratified biomarker modeling in redox biology and aging research^62,63,64^.

These observations highlight the necessity of sex-stratified analysis in redox biology, pointing to the likelihood that biomarker-based assessments require stratification by sex to avoid misinterpretation^65^.

### Factor Analysis and Oxidative Stress Regulation

The use of factor analysis enabled us to move beyond simple pairwise correlations and uncover latent redox networks spanning blood and CSF. This multivariate approach revealed independent biological axes, including CSF-centered oxidative activity, ferric-reducing capacity, and peripheral glutathione signaling. These findings clarify how clusters of oxidative stress biomarkers co-vary in ways that may reflect underlying physiological mechanisms.

Notably, GSH-b was isolated from GSH-csf across all analyses, reinforcing its compartmental specificity and the impact of BBB filtering on glutathione transport^66^. This is consistent with prior studies showing that peripheral glutathione levels do not reliably reflect central depletion^67,68^. Nevertheless, prospective data from the HELIAD study^69^ suggest that higher GSH-b levels may still hold clinical relevance, being linked to lower Alzheimer’s disease risk and attenuated executive decline, even without direct central compartment measurement^70^. By contrast, FRAP spanned blood and CSF compartments, supporting its utility as a systemic marker of antioxidant potential across biological domains.

The variability in factor structures by age and sex further supports the notion that oxidative stress regulation is highly individualized. These results lend support to personalized redox profiling approaches and suggest that a one-size-fits-all biomarker strategy may overlook meaningful subgroup-specific dynamics^71,72^.

### Strengths and Implications for Disease Monitoring

This study offers a comprehensive, multicompartmental view of oxidative stress regulation, integrating blood and CSF biomarker data through both correlation and latent variable modeling. Through subgroup stratification analyses by age and sex, we reveal how oxidative stress coordination might shift across demographic and physiological contexts, potentially uncovering concealed patterns that are not detectable through single-fluid or unstratified approaches.

These findings might carry important implications for clinical biomarker development and disease monitoring. Notably, FRAP emerged as a robust cross-compartment signal, consistently linking systemic and central antioxidant processes across age and sex groups. This consistency positions FRAP as a promising candidate for non-invasive monitoring in clinical trials, longitudinal studies, or routine assessments of redox status. In contrast, biomarkers such as GSH and TAC showed more compartment-specific behavior, indicating that fluid-specific measurements may be necessary for accurate assessment of oxidative balance, particularly in CNS-targeted conditions. Moreover, the observed decline in cross-compartment coordination with age suggests that fixed thresholds for oxidative stress biomarkers may be insufficient. Overall, these results support a more personalized, compartment-aware approach to oxidative stress monitoring in both research and healthcare settings.

### Limitations and Future Directions

As a cross-sectional study, this work cannot support temporal inferences regarding redox trajectory or causal directionality. Longitudinal follow-up is needed to determine whether observed changes in factor structure predict cognitive decline, neurological disease progression, or treatment response. Additionally, the panel of oxidative stress biomarkers used here, while comprehensive, might not capture the full complexity of the redox landscape, including enzymatic antioxidants and reactive oxygen species.

Future work should also address potential circadian effects, which may influence the levels and coordination of redox markers across fluids. Standardized timing of sample collection and inclusion of complementary molecular data (e.g., proteomics, metabolomics, neuroimaging) may further clarify the biological significance of these findings.

## Conclusion

This study presents the first cross-compartment analysis of oxidative stress biomarkers in blood and CSF, revealing that redox regulation is fluid-specific, shaped by age and sex, and only partially coordinated across systems. As individuals age, these networks fragment, and sex-based differences further drive divergent antioxidant strategies. Our findings challenge assumptions about the interchangeability of peripheral and central markers, emphasizing the need for stratified, compartment-aware biomarker models. This approach has important implications for advancing personalized monitoring and therapeutic targeting in aging and neurological disease.

## Methods

### Study Population

This study examines participants from the Aiginition Longitudinal Biomarker Investigation Of Neurodegeneration (ALBION) study, a prospective observational cohort led by the Aiginition University Hospital in Athens, Greece^73, 74^. The ALBION study aims to investigate the biological and clinical markers associated with aging, neurodegeneration, and cognitive decline. The present analysis includes 140 non-demented participants aged 45 years or older. Participants with neurological, psychiatric, or medical conditions associated with a high risk of cognitive impairment or dementia were excluded. These exclusion criteria included but were not limited to Huntington’s disease, multiple sclerosis, Parkinson’s disease, Down syndrome, active alcohol or drug abuse, and major psychiatric conditions such as schizophrenia, bipolar disorder, and major depressive disorder. The ALBION protocol received ethical approval from the Institutional Review Board at Aiginition University Hospital in Athens. Prior to enrollment, all study participants provided written informed consent for their involvement in the study procedures.

### Sample Collection and Processing

Blood and CSF samples were collected at Aiginition University Hospital following standardized protocols. Blood was drawn from the forearm vein, with 5 mL collected into gel-barrier tubes containing a clot activator. Samples were immediately centrifuged (1,370× g for 10 min at 4°C), and the serum was isolated, aliquoted into Eppendorf tubes, and stored at −80°C until analysis. CSF collection followed international guidelines, ensuring standardization of lumbar puncture procedures. Multiple aliquots of CSF were stored at −80°C in polypropylene tubes to prevent degradation.

#### Redox Biomarker Quantification in Serum and CSF

The redox biomarker assays were conducted in blood and CSF using validated spectrophotometric methods.

### Total Antioxidant Capacity (TAC)

TAC was measured based on the method described by Janaszewska and Bartosz^75^. Briefly, 20 μL of serum or CSF was added to 480 μL of 10 mM sodium-potassium phosphate buffer (pH 7.4) and incubated with 500 μL of 0.1 mM 2,2-diphenyl-1-picrylhydrazyl (DPPH) free radical for 1 hour in the dark at room temperature. After centrifugation (15,000 × g, 3 min), the absorbance was recorded at 520 nm, and TAC was expressed as millimoles of reduced DPPH per liter.

### Thiobarbituric Acid-Reactive Substances (TBARS) Assay

Lipid peroxidation levels were assessed by measuring TBARS in serum and CSF. 100 μL of sample was mixed with 500 μL of 35% trichloroacetic acid (TCA) and 500 μL of Tris-HCl buffer and incubated for 10 min at room temperature. One milliliter of 2 M sodium sulfate and 55 mM thiobarbituric acid (TBA) solution was added, and samples were incubated at 95°C for 45 minutes in a boiling water bath. Following cooling at 4°C for 5 min, 1 mL of 70% TCA was added before centrifugation (15,000 × g, 3 min). Absorbance was measured at 530 nm, with TBARS levels expressed using the molar extinction coefficient of malondialdehyde (MDA, 155 × 10^3^ M^−1^ cm^−1^).

### Glutathione (GSH) Assay

GSH levels were determined according to Reddy et al., 2004, with modifications. 100 μL of serum or CSF was mixed with 5% trichloroacetic acid (TCA), centrifuged at 15,000 × g for 5 min at 5°C, and the supernatant was collected. After an additional precipitation step with 5% TCA, the final supernatant was incubated with 700 μL phosphate buffer (400 mM, pH 7.95) and 100 μL DTNB reagent in 40 mM Tris-HCl. After 15 min incubation at room temperature in the dark, absorbance was recorded at 412 nm, and GSH concentration was calculated using the molar extinction coefficient of DTNB (13.6 L/mmol/cm).

### Ferric Reducing Antioxidant Power (FRAP) Assay

Reducing power was measured by the ability of samples to reduce Fe^3+^ to Fe^2+^. 10 μL of serum or CSF was incubated with 240 μL phosphate buffer (0.2 M, pH 6.6) and 250 μL potassium ferricyanide (1% w/v) at 50°C for 20 min. After stopping the reaction with 250 μL of 10% TCA and centrifugation (5,000 × g, 10 min), the supernatant was collected, mixed with 500 μL ferric chloride (0.1% w/v), and incubated at room temperature for 10 min. Absorbance was recorded at 700 nm.

### Hydroxyl Radical Scavenging Activity (HRSA) Assay

HRSA was assessed by measuring hydroxyl radical-induced damage to 2-deoxyribose. 10 μL of serum or CSF was incubated with 225 μL phosphate buffer (0.2 M, pH 7.4), 75 μL of 2-deoxyribose (1 mM), 75 μL of FeSO_4_-EDTA (10 mM), 75 μL of H_2_O_2_ (10 mM), and 290 μL dH_2_O at 37°C for 1 hour. After the incubation, 375 μL of 2.8% TCA and 375 μL of 1% thiobarbituric acid were added, and samples were incubated in a water bath at 95°C for 10 min. Following cooling on ice, centrifugation (3,000 × g, 5 min, 25°C) was performed, and absorbance was monitored at 520 nm.

### Statistical Analyses

All analyses were performed using Python (v3.8) and RStudio (v3.4.3), leveraging packages including NumPy, Pandas, Matplotlib, SciPy, Statsmodels, and statsmodels.stats.multitest for data processing, visualization, and statistical modeling.

Given that most biomarker distributions violated assumptions of normality based on Shapiro-Wilk tests, non-parametric methods were used for bivariate correlations and comparisons. Prior to multivariate modeling, biomarker values were residualized separately using linear regression models, adjusting for relevant covariates (i.e., age or sex depending on the subgroup). For each biomarker, residuals were extracted and z-score standardized. These residuals, not raw biomarker values, were used as inputs for all factor analyses. This ensured that age- or sex-related variance was removed prior to dimensionality reduction, allowing latent factors to reflect covariate-independent variance structures.

Statistical significance was defined at p ≤ 0.05. To account for multiple testing, False Discovery Rate (FDR) correction was applied using the Benjamini-Hochberg procedure to pairwise correlation analyses. Group comparisons by age and sex were reported with unadjusted p-values given the limited number of pre-specified comparisons.

### Blood vs. CSF Antioxidant Biomarker Associations

To examine whether oxidative stress markers in the periphery reflected those in the central nervous system, Spearman’s rank correlation was used to evaluate associations between each blood (-b) and its corresponding CSF (-csf) biomarker. All correlations were adjusted for age and sex to remove potential confounding effects, which was achieved by regressing out these covariates and computing correlations on the residuals. This analysis aimed to assess whether blood biomarkers could indicate oxidative stress within the CNS. FDR correction was applied to account for multiple comparisons.

### Factor Analysis

For the full-sample analysis (Fig. 2), factor analysis was conducted using principal component extraction with varimax rotation (FactorAnalyzer, Python). For age- and sex-stratified analyses (Figs. 3– 4), maximum likelihood factor extraction was used (sklearn.decomposition.FactorAnalysis, scikit-learn) without rotation, adjusting for one covariate within each stratum (age-stratified analyses: adjusted for sex; sex-stratified analyses: adjusted for age). In all analyses, the number of factors was determined using the Kaiser criterion (eigenvalue > 1) and confirmed by scree plot evaluation. For the male subsample, three factors were retained based on the Kaiser criterion; parallel analysis (1,000 permutations, 95th percentile) empirically supported two factors, and the third factor (EV = 1.19) was retained given its theoretical interpretability. Factor loadings were interpreted using a cutoff of ±0.4. The factor structure was evaluated using heatmaps, and the relative contribution of each biomarker to the latent factors was compared across demographic groups to identify key differences in oxidative stress regulation mechanisms.

### Intercorrelations Among Blood Biomarkers

To investigate whether blood oxidative stress markers function as an integrated redox network, pairwise Spearman’s correlations were computed between all blood (_b) biomarkers. Age and sex adjustments were performed using regression residuals to account for potential confounding effects. This analysis allowed for the identification of interdependencies between oxidative stress pathways within the systemic compartment. FDR correction was applied to control for multiple comparisons.

### Intercorrelations Among CSF Biomarkers

Analogous to the blood intercorrelation analysis, Spearman’s correlation was used to assess the relationships between oxidative stress biomarkers within CSF. Given that the CSF provides a more direct reflection of brain-specific oxidative stress processes, this analysis aimed to elucidate how central redox mechanisms interact. Age and sex were adjusted using regression residuals, and FDR correction was applied to minimize false-positive findings.

## Data availability

The raw data in this study can be accessed by contacting the study’s PI. Researchers and interested parties can visit this website to access the data: https://www.maelstrom-research.org/study/albion. For further inquiries about the study, please contact the corresponding author, who can provide additional information and address any specific questions related to the research.

## Acknowledgments

We sincerely thank all the ALBION research participants for their dedicated involvement in this study. This study is supported by the National Network for Research of Neurodegenerative Diseases on the basis of Medical Precision [Grant 2018 E01300001], funded by the General Secretariat of Research and Innovation (GSRI), and by Brain Precision [TAEDR-0535850], funded by the GSRI, through funds provided by the European Union (Next Generation EU) to the National Recovery and Resilience Plan. The funders had no role in study design, data collection, analysis, decision to publish, or manuscript preparation. ANC was supported by a CIHR-Institute of Aging Brain Health and Cognitive Impairment Fellowship [AGE-187963], and acknowledges previous support from the Fonds de Recherche du Québec - Santé (FRQS) Fellowship [335579] and the Réseau Québécoise de recherche sur le vieillissement (RQRV).

## Contributions

A.N.C., S.C., and N.S. conceptualized and designed the study. E.N., E.M. Z.S., and D.K. prepared the biomarker data. A.N.C. and S.C. conducted the data analysis. A.N.C. and S.C. prepared the figures. A.N.C., S.C., and N.S. contributed to the writing and revision of the manuscript, with input from Z.S. and D.K. A.N.C. and N.S. supervised the overall study.

