## Supplemental Materials for "Oxidative Stress Biomarker Profile Dynamics across Blood and Cerebrospinal Fluid"

**Adrián Noriega de la Colina^1,2,3,4^, Zoe Skaperda^5^, Sokratis Charisis^6,7^, Eva Ntanasi^6^, Eirini Mamalaki^6^, Maria Yannakoulia^8^, Christopher** **Papandreou^9,10^, Fotios Tekos^5^, Dimitrios Kouretas^11^, Nikolaos Scarmeas^6,12^.**

^1^Department of Neurology and Neurosurgery, Faculty of Medicine and Health Sciences, McGill University, Montreal, QC, Canada.

^2^ The Montreal Neurological Institute-Hospital, Montreal, QC, Canada.

^3^Department of Mechanical Engineering, Massachusetts Institute of Technology, Cambridge, MA, USA.

^4^Broad Institute of MIT and Harvard, Cambridge, MA, USA.

^5^Department of Biochemistry and Biotechnology, University of Thessaly, Viopolis, Mezourlo, Larissa, Greece

^6^1st Department of Neurology, Aiginition Hospital, National and Kapodistrian University of Athens Medical School, Athens, Greece.

^7^University of Texas Health, San Antonio, TX, USA.

^8^Department of Nutrition and Dietetics, Harokopio University, 17676 Athens, Greece.

^9^Department of Nutrition and Dietetics Sciences, School of Health Sciences, Hellenic Mediterranean University, Siteia, Greece

^10^Institut d'Investigació Sanitària Pere Virgili (IISPV), NeuroÈpia Group, Hospital Universitari Sant Joan de Reus, Reus (Tarragona), Spain

^11^Department of Biochemistry and Biotechnology, University of Thessaly, Viopolis, Mezourlo, Larissa, Greece

^12^Taub Institute for Research in Alzheimer's Disease and the Aging Brain, The Gertrude H. Sergievsky Center, Department of Neurology, Columbia University, New York, New York, USA.

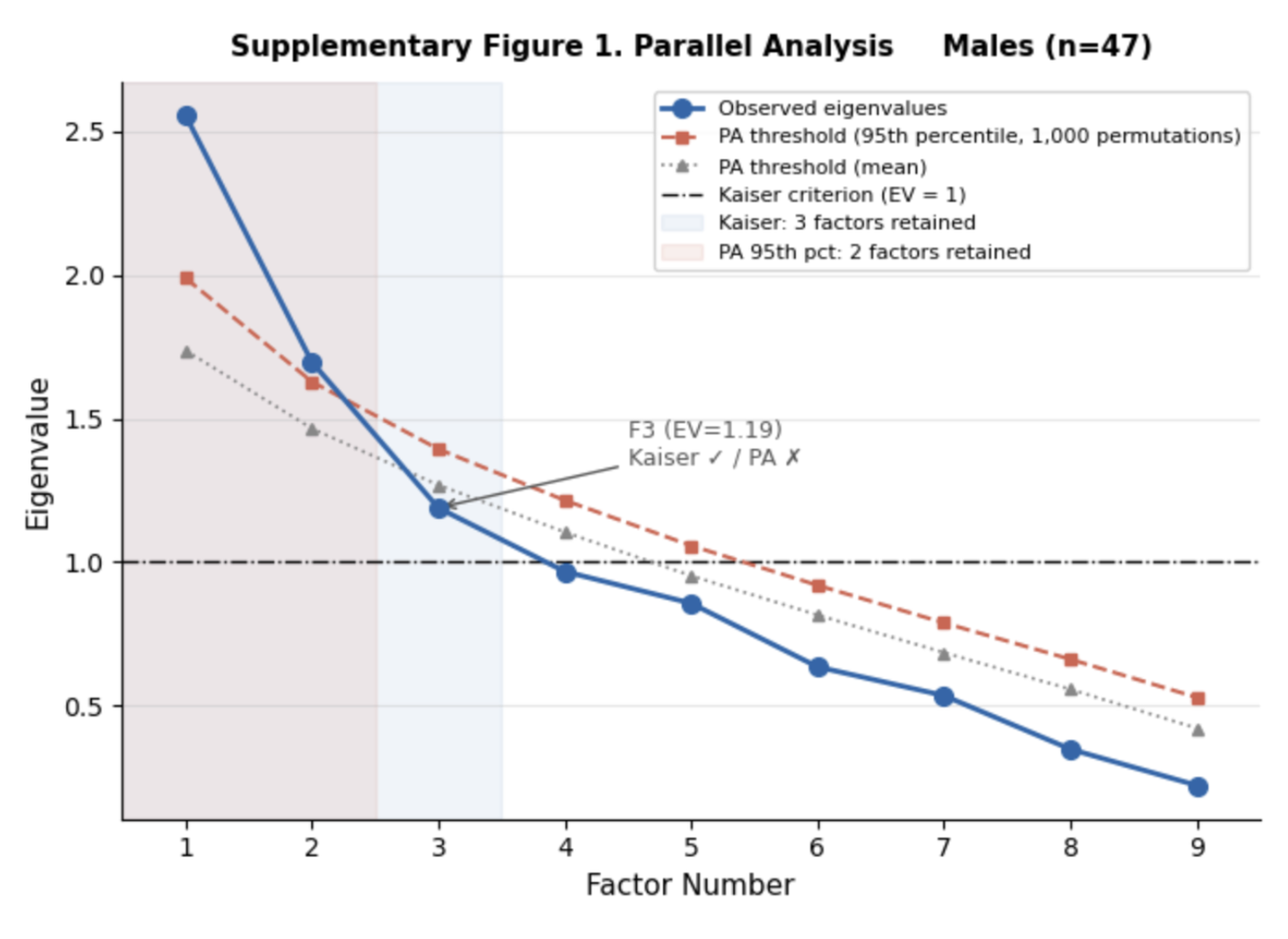

**Supplementary Figure 1. Parallel analysis for factor retention in the male subsample (n=47).**

Scree plot comparing observed eigenvalues (blue circles, solid line) against random-data thresholds generated by parallel analysis (1,000 permutations of matrices with the same dimensions as the male subsample). The red dashed line represents the 95th percentile threshold and the grey dotted line represents the mean threshold across permutations. The dash-dot horizontal line marks the Kaiser criterion (eigenvalue = 1). Blue shading indicates the three factors retained under the Kaiser criterion; pink shading indicates the two factors supported by the 95th percentile parallel analysis threshold. Factor 3 (EV = 1.19) exceeded the Kaiser criterion but fell below the parallel analysis 95th percentile threshold (13.8th percentile of random data), and was retained on theoretical grounds given its interpretable loading pattern. All factor analyses were conducted using maximum likelihood extraction (sklearn.decomposition.FactorAnalysis, scikit-learn), adjusted for age within the male subsample.

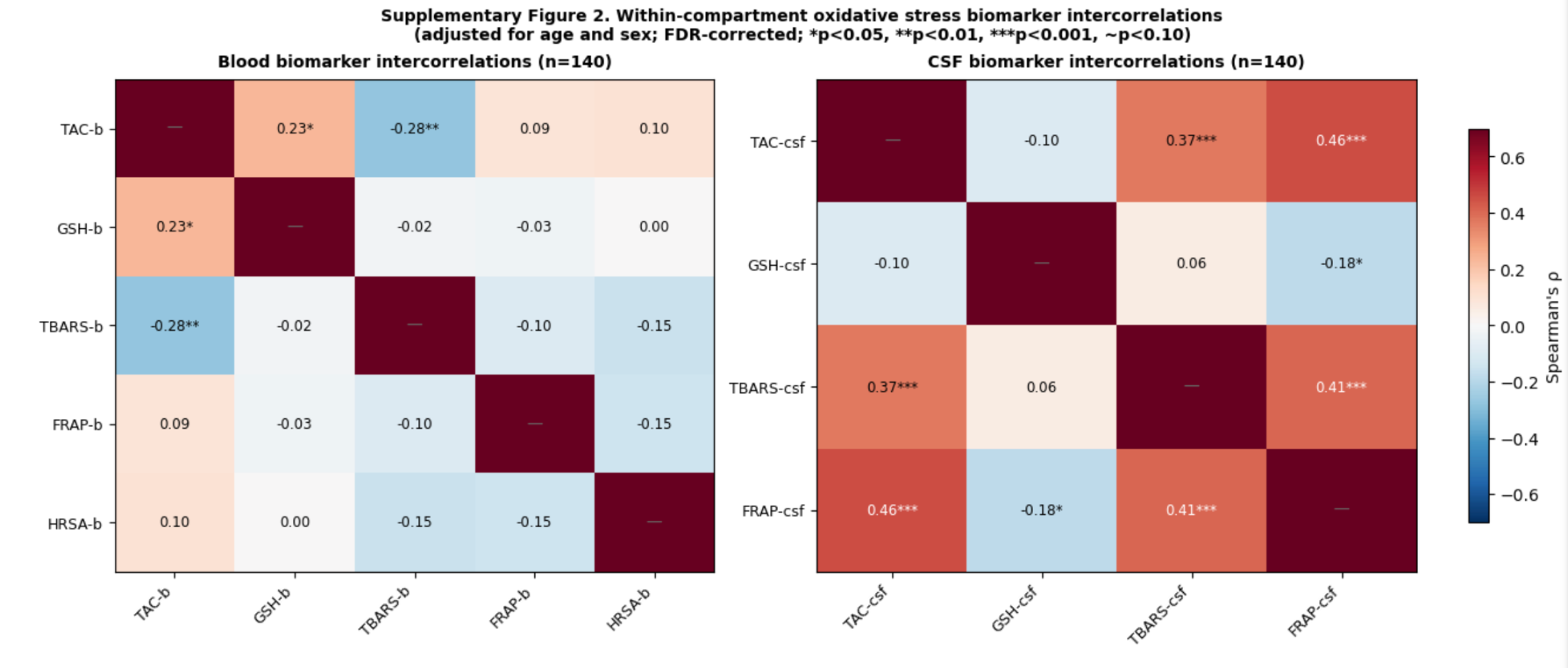

**Supplementary Figure 2. Within-compartment oxidative stress biomarker intercorrelations (n=140).**

Heatmaps display pairwise Spearman correlation coefficients among blood biomarkers (left panel, 5×5) and CSF biomarkers (right panel, 4×4), estimated after adjustment for age and sex by linear regression and z-score standardisation. FDR correction was applied using the Benjamini-Hochberg procedure separately within each compartment. Correlation coefficients are shown in each cell; diagonal cells (—) represent self-correlations. Cell colour encodes the direction and magnitude of the correlation as indicated by the shared colour bar (red = positive, blue = negative). Significance annotations: *p < 0.05, **p < 0.01, ***p < 0.001, ~p < 0.10 (FDR-corrected). Notable within-compartment associations include a positive blood TAC–GSH correlation (ρ = +0.23*) and a negative TAC–TBARS association (ρ = −0.28**), suggesting partial inverse coupling between antioxidant capacity and lipid peroxidation in the peripheral compartment. Within CSF, strong positive associations were observed between TAC-csf and FRAP-csf (ρ = +0.46***) and between TBARS-csf and FRAP-csf (ρ = +0.41***), indicating co-regulated reducing capacity and lipid peroxidation dynamics in the central compartment. Biomarker abbreviations: TAC = Total Antioxidant Capacity; GSH = Glutathione; TBARS = Thiobarbituric Acid-Reactive Substances; FRAP = Ferric Reducing Antioxidant Power; HRSA = Hydroxyl Radical Scavenging Activity.

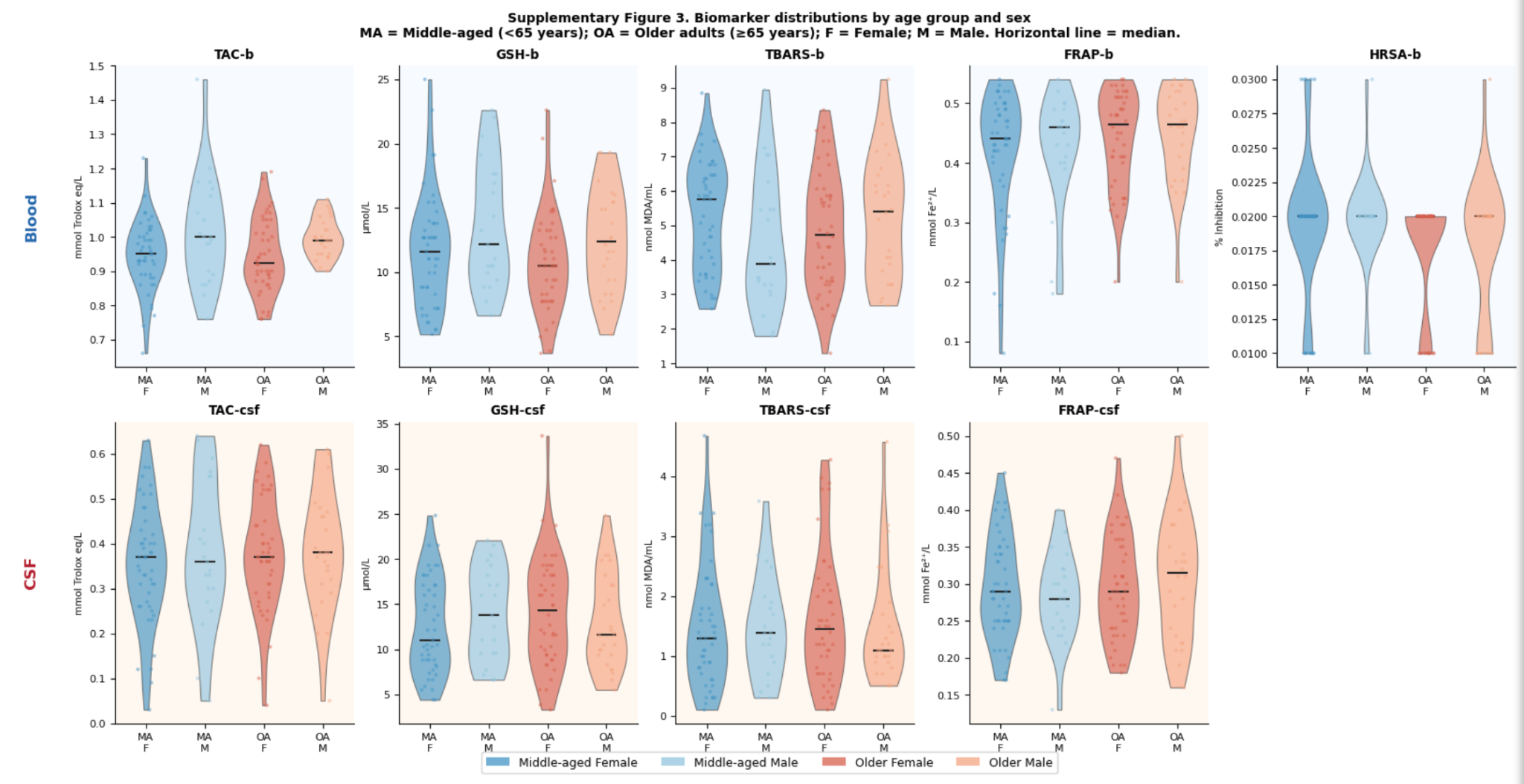

**Supplementary Figure 3. Raw biomarker distributions by age group and sex.**

Violin plots display the distribution of raw (unstandardised) oxidative stress biomarker concentrations across four demographic subgroups: Middle-aged Female (MA F, dark blue), Middle-aged Male (MA M, light blue), Older Female (OA F, dark red/salmon), and Older Male (OA M, light red/peach). Blood biomarkers are shown in the top row (blue background) and CSF biomarkers in the bottom row (orange background). Individual observations are overlaid as jittered dots and the horizontal black line within each violin denotes the group median. Middle-aged is defined as age <65 years (n=68: 47 females, 21 males) and older adults as age ≥65 years (n=72: 46 females, 26 males). The largely overlapping distributions across all four subgroups are consistent with the absence of statistically significant age or sex effects on individual biomarker levels after FDR correction, with the exception of TAC-b and GSH-b (higher in males; reported in Results). The wide within-group variance visible across all panels, particularly for GSH-b, TBARS-csf, and GSH-csf, underscores the limited discriminative utility of individual biomarker levels and motivates the latent variable approach adopted in the main analyses. Biomarker abbreviations and units: TAC = Total Antioxidant Capacity (mmol Trolox eq/L); GSH = Glutathione (μmol/L); TBARS = Thiobarbituric Acid-Reactive Substances (nmol MDA/mL); FRAP = Ferric Reducing Antioxidant Power (mmol Fe²⁺/L); HRSA = Hydroxyl Radical Scavenging Activity (% inhibition; blood only).
